# Performance of a Self-Supervised Pretrained Neural Network for Orthopedic Radiograph Classification

**DOI:** 10.64898/2026.08.07.26359986

**Authors:** Rohan Bagchi, Nicholas J. Yee, John Y. Kwon, Atta Taseh, Soheil Ashkani-Esfahani

## Abstract

**Purpose:** To evaluate whether domain-adaptive self-supervised pretraining on musculoskeletal radiographs improves fracture classification and attribution faithfulness relative to ImageNet-pretrained baselines.

**Materials and Methods:** This study (June 2025–May 2026) used previously acquired radiographs to compare three ResNet-50 initializations: supervised ImageNet pretraining (control), self-supervised ImageNet pretraining (DINO), and DINO with additional domain-adapted pretraining on 44,029 musculoskeletal radiographs (DINO-Ortho). All models underwent supervised fine-tuning in three experiments: in-distribution (MURA and FracAtlas datasets), out-of-distribution (an external dataset of 5,365 calcaneal radiographs from 1,775 patients), and initial weights (calcaneal radiographs only). Metrics included sensitivity, specificity, test accuracy, area under the receiver operating characteristic curve (AUROC), and Cohen’s kappa; attribution faithfulness was quantified using Remove and Debias scores from Grad-CAM saliency maps.

Comparisons used DeLong and Friedman tests.

**Results:** Classification performance did not differ significantly between DINO-Ortho and either baseline in any experiment (DINO-Ortho AUROC, 0.89 in-distribution and 0.95 with initial weights). All three models discriminated poorly out-of-distribution (control, 0.59; DINO, 0.57; DINO-Ortho, 0.58). DINO-Ortho showed significantly higher attribution faithfulness than both baselines in all three experiments, including out-of-distribution (25.39 vs −10.41 and 2.14; *P* < .001) and initial weights (20.88 vs 11.51 and 1.27; *P* < .001). Qualitative rankings favored DINO-Ortho but did not differ significantly.

**Conclusion:** Domain-adapted self-supervised pretraining on musculoskeletal radiographs improved attribution faithfulness while maintaining classification performance comparable to ImageNet-pretrained baselines; no model generalized adequately to external radiographs without task-specific fine-tuning.

## Introduction

Musculoskeletal (MSK) radiographs are the most commonly ordered imaging study in orthopedics and serve as the first-line imaging modality for fracture diagnosis (1,2). Interpretation can be challenging, however, because fractures may be subtle and are often read in high-pressure settings such as the emergency department (2–4). Furthermore, initial interpretation is often performed by clinicians without subspecialty MSK imaging training. This can lead to missed fracture diagnoses and delayed treatments (2–4). Deep learning models have shown promise for fracture detection and classification on radiographs (5,6). Clinical adoption depends not only on accuracy but on whether predictions are interpretable, particularly when a model may base its prediction on image features unrelated to the underlying pathology (7). Predictions should therefore be driven by clinically relevant anatomy rather than background features or spurious correlations (8–10).

Image classification models are traditionally trained with supervised learning (SL), which learns directly from expert-labeled examples and performs well when annotated data are available (11). In medical imaging, however, expert annotation is costly and labeled datasets are often small (11,12). SL models are also prone to overfitting, generalizing poorly to unseen data because they rely on training-specific features (12). Self-supervised learning (SSL) offers an alternative when unlabeled images are abundant (13). SSL derives supervisory signals from the images themselves, for example by comparing differently augmented views of the same image (13). Previous literature suggests that SSL models can learn meaningful feature representations that improve model explainability (14).

A prominent approach to SSL follows Meta’s Distillation with No Labels (DINO) algorithm, which uses teacher-student self-distillation to learn visual representations without manual labels (15). DINOv1 uses a convolutional neural network backbone trained on ImageNet-1k and learns general visual features that transfer across downstream imaging tasks (15). ImageNet contains few, if any, diagnostic radiographs, so ImageNet-pretrained models may not represent the anatomy, contrast, and pathology patterns of MSK radiographs. Most deep learning models nonetheless still initialize from these generic weights (11,12).

Continuing SSL pretraining on radiographs could align these representations with MSK anatomy and yield more clinically meaningful attributions. In MSK imaging, it remains unclear whether domain-adaptive SSL pretraining improves classification performance, external-domain performance, or attribution faithfulness compared with generic ImageNet-supervised or ImageNet-self-supervised initialization.

This study developed and evaluated DINO-Ortho, an MSK-domain-adapted ResNet-50 created by further self-supervised pretraining of DINOv1 on MSK radiographs, to determine whether domain-adaptive self-supervision could provide more robust and explainable image representations than generic SL or SSL models. We hypothesized that an MSK-specific pretrained model could serve as an effective initialization for downstream MSK fracture detection tasks, achieving superior diagnostic performance and attribution faithfulness.

## Materials and Methods

### Study Design and Datasets

This study was conducted using previously acquired radiographs at a local tertiary healthcare institution. Institutional review board (IRB) approval was obtained for use of the institutional dataset of calcaneal radiographs, with waiver of informed consent because of the retrospective study design (IRB number withheld for anonymization). This study was designed and reported in accordance with the Checklist for Artificial Intelligence in Medical Imaging guidelines (16).

The Musculoskeletal Radiographs (MURA) dataset was an upper extremity radiograph dataset containing 40,005 radiographs (23,602 normal, 16,403 abnormal), each labeled as normal or abnormal (17). FracAtlas contained 4,024 radiographs of the hand, leg, hip, and shoulder collected from three hospitals (3,307 normal, 717 fractured) (18). A calcaneus dataset from a single hospital system containing 5,365 radiographs from 1,775 patients (551 with calcaneus fracture, 1,224 without; up to five views per patient) served as the institutional dataset. Series were included if they comprised complete anteroposterior, oblique, and lateral foot views acquired during routine clinical care between 2015 and 2022. No standardized research acquisition protocol was applied. Imaging data were extracted from the institutional imaging archive in anonymized form. MURA and FracAtlas were publicly released, de-identified datasets. Calcaneus labels were derived from the clinical radiology reports of board-certified diagnostic radiologists, whereas the labels released with MURA and FracAtlas were used without modification. No images were re-read for this study.

MURA used its predefined training split, with the predefined validation split divided 70:30 into validation and test data. FracAtlas was split 70:15:15 for train:validation:test; both were split at the image level, not by patient identifier. The calcaneus dataset was split 70:15:15 at the patient level.

### Model Initialization Strategies

Three models with a ResNet-50 backbone were compared in this study (Table 1). The control model was a standard ResNet-50 model pretrained on ImageNet using SL. The DINO model was a DINOv1 ResNet-50 model pretrained on ImageNet using SSL (15). DINO-Ortho extended this DINOv1 model with additional SSL pretraining on the MURA and FracAtlas datasets. Although both are labeled datasets, all annotations were ignored during the SSL step and images were treated as unlabeled.

**Table 1.** Summary of experimental design and dataset allocation.

| Experiment | Supervised fine-tuning data | Test data | Purpose |
| --- | --- | --- | --- |
| In-distribution | MURA + FracAtlas<br>train/validation split | Held-out MURA +<br>FracAtlas test split | Internal validity |
| Out-of-distribution | MURA + FracAtlas<br>train/validation split | Full calcaneus dataset | External validity |
| Initial weights | Calcaneus train/validation split | Held-out calcaneus test<br>split | Effect of initialization strategy |

### DINO-Ortho Pretraining Configuration

A major modification introduced in DINO-Ortho was the replacement of the multi-crop (global–local) random cropping that was used in the original DINO algorithm with anatomically guided cropping. Random cropping generated views of arbitrary image regions, which in MSK radiographs risks removing part of a bone or fracture site. To address this, for images demonstrating fracture in the FracAtlas dataset, bounding box annotations on each image were used to generate cropped views centered on the fracture region (18). Each bounding box encompassed the fracture site, with 60-pixel padding applied on each image to preserve spatial context. Cropping was not applied to normal FracAtlas or MURA images. All images underwent DINO-framework augmentations, including random rotations, geometric distortions, and color jittering (15).

The DINO-Ortho SSL step used a student–teacher architecture with a ResNet-50 backbone producing 2048-dimensional feature representations (15). All experiments used a fixed random seed for reproducibility. A full list of hyperparameters can be seen in Supplementary Table 1 and Supplementary Table 2. The final DINO-Ortho model was selected at the epoch with the lowest SSL training loss, because no labels or held-out validation sets were available during self-supervised pretraining.

### Supervised Fine-Tuning Experiments

After pretraining, the three models were evaluated through three downstream fine-tuning experiments (Table 1). Hyperparameters were optimized based on downstream validation performance and were held constant across all three models (Supplementary Table 2). Bounding-box cropping was applied only during self-supervised pretraining; supervised fine-tuning used uncropped images for all three models. The model weights from each experiment yielding the highest validation accuracy were evaluated on test data.

In the first experiment, termed “in-distribution,” the three models were fine-tuned on MURA and FracAtlas and tested on held-out data from those same datasets. In the second experiment, termed “out-of-distribution,” the models were fine-tuned on MURA and FracAtlas and tested on the unseen external calcaneus dataset. In the third experiment, termed “initial weights,” the models were fine-tuned on the calcaneus dataset and tested on a held-out portion of that dataset.

### Explainability and Attribution Faithfulness

Model explainability was evaluated using Gradient-weighted Class Activation Mapping (Grad-CAM), computed at the first convolutional layer of layer4, the final residual block of the ResNet-50 backbone (19). To quantitatively assess attribution faithfulness, a combined Remove and Debias (ROAD) calculation was performed (20). The combined ROAD score was calculated as (LeRF – MoRF)/2, where LeRF and MoRF were the Least Relevant First and Most Relevant First perturbation strategies, respectively (20). For MoRF, the most activated Grad-CAM regions were removed in 10% increments from 10% to 50% of pixels, and the mean drop in predicted probability for the fractured class was recorded. LeRF was calculated identically but removed the least activated regions; a larger drop under MoRF and a smaller drop under LeRF indicated greater reliance on a specific region. ROAD score analysis was restricted to fractured images correctly classified by all three models to ensure paired comparison across the same image set. To ensure consistency, Grad-CAM saliency maps and ROAD scores were computed with respect to the fractured class rather than the predicted class.

In addition to the quantitative ROAD analysis, a qualitative visual review of Grad-CAM saliency maps was performed by two investigators on 50 randomly sampled fractured calcaneus images for the initial weights experiment test set. The raters were blinded to the model used to generate each saliency map and the model predictions. For each image, the raters were given three saliency maps and ranked them as best (score = 1), second best (score = 2), and worst (score = 3). Maps were ranked on two criteria considered together: the extent of background highlighting (minimal or none, some, or more background than anatomy) and localization to the fracture (fracture region only, fracture region plus other areas, or fracture region missed entirely). The map with the least background highlighting and most focused localization was ranked first; ties were not permitted. Then, average scores per model were computed. Inter-rater and intra-rater reliability were quantified using quadratic weighted Cohen’s kappa and interpreted using the Landis and Koch scale as slight (κ ≤ 0.20), fair (0.21–0.40), moderate (0.41–0.60), substantial (0.61– 0.80), or almost perfect (0.81–1.00) (21).

### Performance Metrics and Statistical Analysis

Model performance was evaluated using sensitivity, specificity, test accuracy, AUROC, and Cohen’s kappa (16). Confidence intervals for all reported metrics were calculated by bootstrapping with replacement over 2,000 iterations and reported at the 95% level. Statistical pairwise comparisons of AUROC across the three models were performed using DeLong tests with Bonferroni correction. Classification accuracy between the in-distribution and out-of-distribution experiments was compared for each model using a two-proportion z test. ROAD scores were compared across models using the Friedman test with post-hoc Wilcoxon signed-rank tests with Bonferroni correction. All statistical tests used a significance level α = 0.05. No a priori sample size calculation was performed; all datasets had fixed, predetermined sample sizes. Analyses, model training, and inference used Python 3.11.7, PyTorch 2.5.1, and an NVIDIA GeForce RTX 4090 GPU.

## Results

### Patient and Dataset Demographics

The MURA dataset contained no demographic information, as it was de-identified for patient privacy (17). The FracAtlas dataset contained radiographs from patients aged 8 months to 78 years old and was 62% (2,496/4,024) male and 38% (1,528/4,024) female (18). Males accounted for 85.4% (612/717) of the fracture cases. For the calcaneus dataset, females represented 43% (763/1,775) of the patients (mean age 60 ± 17 years) and males represented 57% (1,012/1,775) (mean age 53 ± 15 years).

### In-Distribution Experiment

The in-distribution experiment used a held-out set of 1,565 radiographs (1,004 normal, 561 abnormal) from MURA and FracAtlas. Classification performance was similar across models, with AUROC values of 0.89 and test accuracy of 0.84 for all three models and no significant pairwise AUROC differences (Table 2, Supplementary Table 3). Pairwise AUROC comparisons for this experiment are shown in Table 3. DINO-Ortho had the highest ROAD score, which was significantly higher than both control and DINO (21.09 vs −8.76 and 2.04, respectively; overall P < .001; all pairwise P < .001; χ² = 117.07; Table 4). Figure 1 shows an example lower extremity fracture from the in-distribution experiment.

**Table 2.** Performance of fine-tuned models across all three experiments: A) In-distribution, B) Out-of-distribution, and C) Initial weights. Values are reported with 95% confidence intervals in parentheses. Bolded text demonstrates the highest performance.

| Model | Sensitivity | Specificity | AUROC | Cohen's kappa | Test Accuracy |
| --- | --- | --- | --- | --- | --- |
| **A.In-distribution** |  |  |  |  |  |
| Control | 0.66 (0.62–0.70) | **0.94 (0.93–0.96)** | **0.89 (0.87–0.90)** | **0.64 (0.60–0.68)** | **0.84 (0.82–0.86)** |
| DINO | **0.68 (0.64–0.72)** | 0.92 (0.90–0.94) | **0.89 (0.87–0.90)** | 0.63 (0.59–0.67) | **0.84 (0.82–0.86)** |
| DINO-Ortho | 0.67 (0.63–0.71) | 0.93 (0.91–0.95) | **0.89 (0.87–0.91)** | 0.63 (0.59–0.67) | **0.84 (0.82–0.86)** |
| **B.Out-of-distribution** |  |  |  |  |  |
| Control | 0.42 (0.40–0.44) | **0.77 (0.75–0.78)** | **0.59 (0.57–0.61)** | **0.19 (0.16–0.21)** | **0.66 (0.64–0.67)** |
| DINO | 0.35 (0.33–0.38) | **0.77 (0.76–0.78)** | 0.57 (0.56–0.59) | 0.13 (0.10–0.16) | 0.64 (0.63–0.65) |
| DINO-Ortho | **0.47 (0.44–0.49)** | 0.66 (0.64–0.67) | 0.58 (0.56–0.59) | 0.12 (0.09–0.15) | 0.60 (0.59–0.61) |
| **C.Initial weights** |  |  |  |  |  |
| Control | **0.86 (0.81–0.90)** | 0.92 (0.89–0.94) | 0.93 (0.91–0.95) | 0.77 (0.72–0.82) | 0.90 (0.87–0.92) |
| DINO | 0.85 (0.80–0.89) | **0.94 (0.92–0.96)** | **0.95 (0.94–0.97)** | **0.80 (0.75–0.84)** | **0.91 (0.89–0.93)** |
| DINO-Ortho | 0.85 (0.81–0.89) | 0.93 (0.90–0.95) | **0.95 (0.93–0.96)** | 0.78 (0.74–0.83) | 0.90 (0.88–0.92) |

*AUROC = area under the receiver-operating characteristic curve*

**Table 3.** DeLong pairwise AUROC comparison P-values.

| Comparison | In-distribution | Out-of-distribution | Initial weights |
| --- | --- | --- | --- |
| Control vs DINO | >.99 | .04* | .044* |
| Control vs DINO-Ortho | >.99 | .16 | .27 |
| DINO-Ortho vs DINO | >.99 | >.99 | .53 |

\* Denotes significance
*Control = ResNet-50 with supervised ImageNet pretraining; DINO = ResNet-50 with self-supervised ImageNet pretraining; DINO-Ortho = DINO with additional self-supervised pretraining on musculoskeletal radiographs.*

**Table 4.** Mean ROAD scores (percentage points) on fracture predictions across experiments (N = 155, 67, and 223 images for the in-distribution, out-of-distribution, and initial weights experiments, respectively, in which all three models were correct), with pairwise post-hoc comparisons (Bonferroni-corrected P values) for all three experiments. Bolded text demonstrates the highest ROAD score per experiment.

| Model | In-distribution | Out-of-distribution | Initial weights |
| --- | --- | --- | --- |
| Control | -8.76 | -10.41 | 11.51 |
| DINO | 2.04 | 2.14 | 1.27 |
| DINO-Ortho | **21.09** | **25.39** | **20.88** |
| $\chi^2$ statistic* | 117.07 | 64.57 | 109.52 |
| $P$-value* | <.001 | <.001 | <.001 |
| **Pairwise comparisons** |  |  |  |
| **(corrected $P$-values)** |  |  |  |
| Control vs DINO | <.001 | <.001 | <.001 |
| Control vs DINO-Ortho | <.001 | <.001 | <.001 |
| DINO vs DINO-Ortho | <.001 | <.001 | <.001 |

*ROAD = Remove and Debias*
*\*From Friedman test*

**Figure 1.**
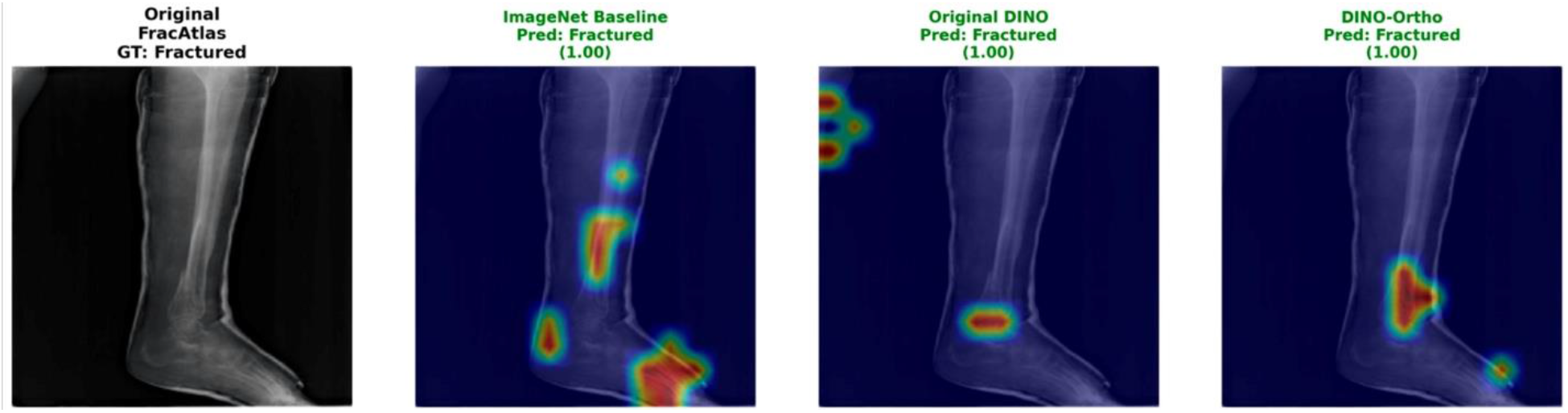
Grad-CAM visualization for a lower extremity radiograph from the FracAtlas dataset in the in-distribution experiment. From left to right: original radiograph (ground truth: fracture at distal tibia), ImageNet-pretrained control, original DINO, and DINO-Ortho. All models correctly predict a fractured image. Saliency maps illustrate regions contributing most strongly to the prediction, with warmer colors indicating higher importance. Numbers in parentheses represent model confidence, ranging from 0 (lowest confidence) to 1 (highest confidence). The control model attributed its prediction to non-pathologic regions including the tibial shaft, calcaneus, and midfoot and missed the clinically relevant anatomy, whereas DINO localized the fracture but also highlighted radiolucent background regions. DINO-Ortho primarily highlighted the fracture region.

### Out-of-Distribution Experiment

The out-of-distribution experiment used a test set of 5,365 radiographs from 1,775 patients (3,671 normal, 1,694 abnormal) from the external calcaneus dataset. Classification performance decreased significantly for all models compared with the in-distribution experiment (two-proportion z test, P < .001 for each model). AUROC was low and similar across models, at 0.59 for control, 0.57 for DINO, and 0.58 for DINO-Ortho (Table 2, Supplementary Table 3). Only the control versus DINO comparison differed significantly (P = .04; Table 3). DINO-Ortho had the highest ROAD score, which was significantly higher than both control and DINO (25.39 vs −10.41 and 2.14, respectively; overall P < .001; all pairwise P < .001; χ² = 64.57; Table 4). Figure 2 shows an example calcaneal fracture from the out-of-distribution experiment.

**Figure 2.**
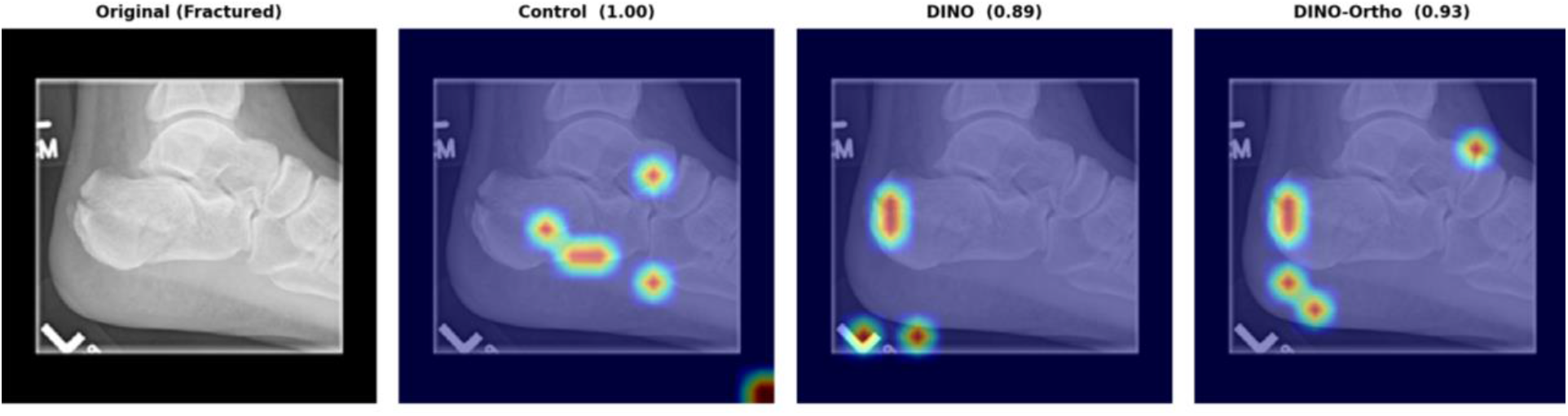
Grad-CAM visualization for a foot and ankle radiograph from the external calcaneus dataset in the out-of-distribution experiment. From left to right: original radiograph (ground truth: fracture at posterior calcaneus), ImageNet-pretrained control, original DINO, and DINO-Ortho. All models correctly predict a fractured image. Saliency maps illustrate regions contributing most strongly to the prediction, with warmer colors indicating higher importance. Numbers in parentheses represent model confidence, ranging from 0 (lowest confidence) to 1 (highest confidence). The control and DINO models attended to background regions, and the control model also missed the fracture; DINO-Ortho was the only model to both highlight the fracture region and avoid background.

### Initial Weights Experiment

The initial weights experiment used 814 radiographs (534 normal, 280 abnormal) from the external calcaneus dataset. All models achieved high classification performance, with AUROC values of 0.93 for control, 0.95 for DINO, and 0.95 for DINO-Ortho (Table 2, Supplementary Table 3). Pairwise AUROC comparisons for this experiment are shown in Table 3. DINO demonstrated a significantly higher AUROC compared with control (0.95 vs 0.93; P = .044; Table 3). DINO-Ortho had the highest ROAD score, which was significantly higher than both control and DINO (20.88 vs 11.51 and 1.27, respectively; overall P < .001; all pairwise P < .001; χ² = 109.52; Table 4).

DINO-Ortho demonstrated the best (lowest) mean qualitative ranking, although rankings did not differ significantly among models (1.89 vs 2.20 and 1.92, respectively; P = .32; χ² = 2.31; Supplementary Table 4). Intra-rater agreement was almost perfect (*κ **=*** 0.84 and 0.88 for Raters 1 and 2, respectively, Supplementary Table 5) and inter-rater agreement was substantial (κ = 0.72, Supplementary Table 5) for this ranking.

Figure 3 shows an example calcaneal fracture from the initial weights experiment. A single calcaneal radiograph is shown with saliency maps from both the out-of-distribution and initial weights experiments (Figure 4).

**Figure 3.**
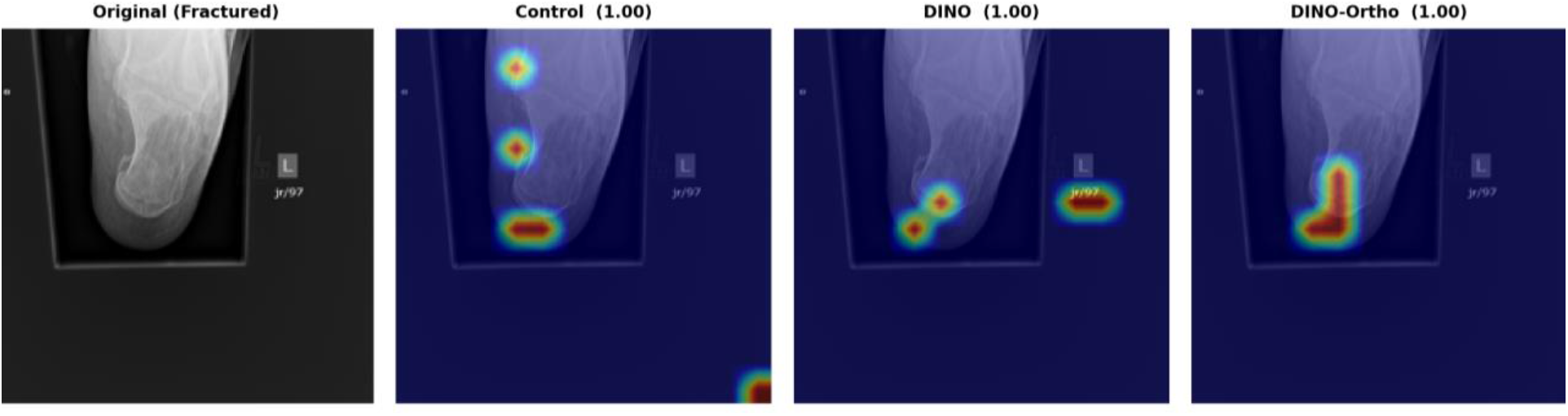
Grad-CAM visualization for a foot and ankle radiograph from the external calcaneus dataset in the initial weights experiment. From left to right: original radiograph (ground truth: calcaneal fracture), ImageNet-pretrained control, original DINO, and DINO-Ortho. All models correctly predict a fractured image. Saliency maps illustrate regions contributing most strongly to the prediction, with warmer colors indicating higher importance. Numbers in parentheses represent model confidence, ranging from 0 (lowest confidence) to 1 (highest confidence). DINO-Ortho localized to the posterior and plantar calcaneus with residual activation along the collimation edge, whereas the control model activated over an image corner, and DINO highlighted over the radiographic side marker.

**Figure 4.**
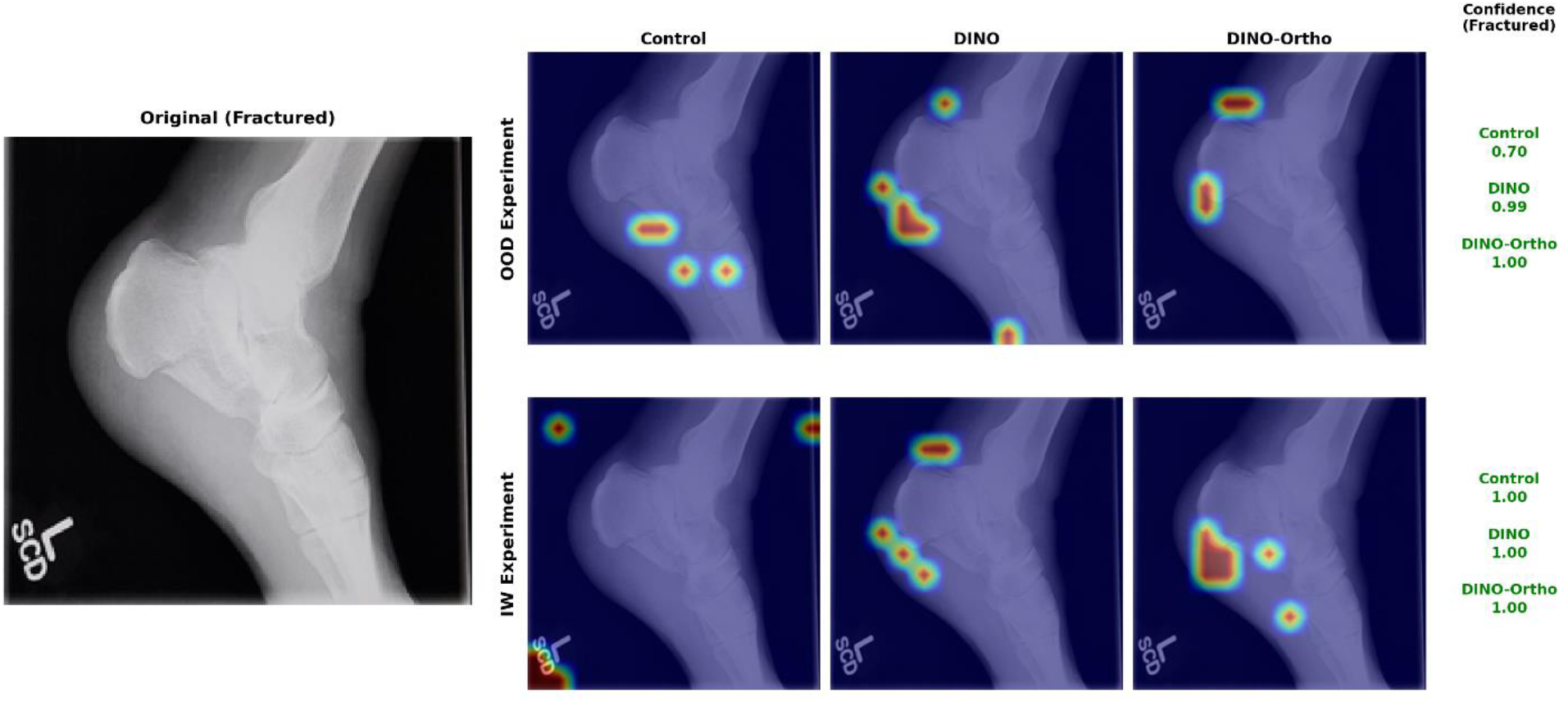
Grad-CAM visualization for a foot and ankle radiograph from the external calcaneus dataset from two experiments. From left to right: original radiograph (ground truth: calcaneal fracture), ImageNet-pretrained control, original DINO, and DINO-Ortho. All models correctly predict a fractured image. Saliency maps illustrate regions contributing most strongly to the prediction, with warmer colors indicating higher importance. Numbers in parentheses represent model confidence, ranging from 0 (lowest confidence) to 1 (highest confidence). In both experiments, DINO-Ortho’s saliency maps highlighted less background than those of the control model, which activated over image corners and the radiographic marker; differences from DINO were less pronounced. *OOD: Out-of-distribution; IW: Initial weights*

## Discussion

In this study, we developed DINO-Ortho, an MSK domain-adapted model created by additional SSL pretraining of DINOv1 on 44,029 MSK radiographs. DINO-Ortho achieved fracture-classification performance comparable to ImageNet-pretrained baselines in all three experiments, although all three models discriminated poorly out-of-distribution (AUROC 0.57–0.59). These outcomes showed that domain-adaptive SSL pretraining did not improve classification accuracy over general-image pretraining but did improve the faithfulness of model attributions in all three experiments.

These findings were consistent with prior reports that domain-specific representations alter where saliency methods focus attention in medical imaging. Adebayo et al. demonstrated that many saliency methods, including Grad-CAM, can produce visually plausible maps even when model parameters are randomized, motivating quantitative perturbation-based metrics such as ROAD to verify whether attribution reflects what the model uses (20,22). Suara et al. similarly reported that Grad-CAM often highlights non-diagnostic regions across medical imaging datasets, particularly with natural-image pretraining alone, and recommended pairing qualitative inspection with quantitative faithfulness evaluation (23). In our study, ImageNet-pretrained models classified well in the in-distribution and initial weights experiments but localized to background or radiolucent regions, whereas DINO-Ortho performed comparably with higher ROAD scores. That discrepancy is consistent with this prior observation: classification accuracy alone is insufficient to characterize model explainability on MSK radiographs. Notably, control ROAD scores were negative in-distribution and out-of-distribution (−8.76 and −10.41): removing the regions its saliency maps ranked least relevant degraded predictions more than removing those ranked most relevant, indicating attribution anticorrelated with the features the model used.

The improvement in explanation faithfulness is likely due to the added SSL pretraining on MSK radiographs. While previous studies focused on SL fine-tuning an SSL model such as DINO, DINO-Ortho used an additional SSL step on MSK radiographs before SL fine-tuning. This approach could extend to other imaging modalities or organ systems and may be more feasible than assembling large labeled datasets.

Domain-adaptive SSL pretraining has shown similar benefits in other medical imaging domains. Azizi et al. found that SSL pretraining improved downstream chest radiograph and dermatology classification over ImageNet-supervised initialization, particularly with limited labels (24). Beyond radiology, Wang et al. found that SSL pretraining outperformed ImageNet-pretrained baselines on histopathologic whole-slide image classification (25). In ophthalmology, Holmberg et al. found that a self-supervised retinal-thickness prediction task improved diabetic retinopathy classification (26). Consistent with these reports, an additional SSL step on MSK images was sufficient to alter where the model attends.

Generalizability was evaluated on an unseen, single-institution calcaneus dataset. Although all three models discriminated poorly on out-of-distribution calcaneus radiographs, with no significant difference between DINO-Ortho and either baseline, DINO-Ortho demonstrated higher ROAD-based attribution faithfulness. Out-of-distribution, DINO-Ortho had the highest sensitivity but the lowest specificity and accuracy, indicating a tendency to classify radiographs as fractured rather than improved discrimination. Qualitative review of saliency maps from the initial weights experiment suggested DINO-Ortho highlighted less background and focused more on fracture regions than the ImageNet-pretrained baselines. Together, these findings indicate that domain-adaptive pretraining improved attribution quality without improving generalization to an unseen anatomic region.

Several factors likely contributed to the uniform decline in out-of-distribution performance. The calcaneus is absent from both training datasets: MURA comprises upper extremity radiographs, and FracAtlas covers the hand, leg, hip, and shoulder. The out-of-distribution experiment therefore asked the models to classify an unfamiliar anatomic region and view; differences in acquisition and positioning would further compound this difference. Zech et al. similarly found that a pneumonia-detection model performed worse on external radiographs, attributing part of the effect to site-specific features rather than pathology (9). That all three initializations degraded together, and improved to AUROC 0.93–0.95 once fine-tuned on calcaneus data, suggests the limitation reflects domain gap rather than pretraining strategy; generalizability cannot be established from a single external dataset.

This study had several limitations. First, out-of-distribution testing was performed on a single external dataset limited to a binary fracture-classification task in the calcaneus, an anatomically simple region relative to sites such as the wrist, hand, or axial skeleton, so results may not generalize to more complex anatomy, other pathologies, or other tasks such as implant detection or segmentation. Second, although class labels were withheld during SSL pretraining, FracAtlas bounding box annotations guided anatomically focused crop augmentation for fractured images, a mild form of spatial supervision that limits the claim of fully label-free pretraining; this could be resolved in future work with standard random cropping or region proposals from an unsupervised detector. Third, ROAD captures internal consistency rather than anatomical correctness, so higher scores do not always imply anatomically meaningful attribution (20,22) although a high ROAD score can support consistency for a Grad-CAM map that already highlights relevant regions.

In conclusion, domain-adaptive SSL pretraining improved ROAD-based attribution faithfulness while maintaining classification performance comparable to ImageNet-pretrained initialization in all three experiments. None of the three initializations discriminated adequately on external radiographs without task-specific fine-tuning. Domain-adapted models should therefore be considered for MSK radiograph classification when fine-tuning on the target task is feasible. Future studies should evaluate domain-adaptive pretraining across multiple institutions and anatomic regions and on other tasks such as segmentation and implant detection.

## Data Availability

All data produced in the present study are available upon reasonable request to the authors.

## Acknowledgements

The authors gratefully acknowledge the patients whose medical records and data made this study possible. We would also like to thank the FARIL-MGB lab for all their support. This study did not receive any financial support from any entity.

## Data sharing statement

Data generated or analyzed during the study are available from the corresponding author by request. The institutional calcaneus radiograph dataset is proprietary to Mass General Brigham and cannot be shared publicly; deidentified imaging data and fracture labels may be requested from the corresponding author upon publication, subject to a data use agreement and institutional review board approval at the requesting institution, with no predetermined end date for availability. The publicly available MURA (doi.org/10.48550/arXiv.1712.06957) and FracAtlas (doi.org/10.1038/s41597-023-02432-4) datasets were analyzed during the study and are available from their original sources without restriction. The DINO-Ortho pretrained model weights and inference code will be deposited in a public repository upon acceptance and will remain openly available without restriction.

## Supplementary Information

**Supplementary Table 1.** Hyperparameters, self-supervised learning.

| Hyperparameter | DINOv1 | DINO-Ortho |
| --- | --- | --- |
| Backbone | ResNet-50 | ResNet-50 |
| Projection head layers | 3 (MLP) | 3 (MLP) |
| Hidden dimension | 2048 | 1024 |
| Projection dimension | 256 | 256 |
| Prototypes | 65536 | 512 |
| Student temperature | 0.1 | 0.12 |
| Warmup teacher temperature | 0.04 | 0.04 |
| Final teacher temperature | 0.04 | 0.04 |
| Teacher temperature warmup epochs | 0 | 30 |
| Center momentum | 0.9 | 0.95 |
| EMA momentum schedule | Cosine, 0.996 → 1.0 | 0.95 → 0.98 → 0.995 |
| Optimizer | AdamW | AdamW |
| Base learning rate | 0.0005 | 0.0001 |
| Weight decay | 0.04 | 0.01 |
| Gradient clipping | 3.0 | 0.5 |
| Batch size | 64 | 64 |
| Image size | 224×224 | 224×224 |
| Maximum epochs | 100 | 500 |
| Early stopping patience | None | 30 |
| Best model selection | Final epoch | Lowest training loss |

*EMA: Exponential Moving Average*
*MLP: Multilayer Perceptron*

**Supplementary Table 2.** Hyperparameters, supervised learning, all three models.

| Hyperparameter | Value |
| --- | --- |
| Backbone | ResNet-50 |
| Pre-training | Model dependent (see Methods section) |
| Classifier | Linear, 2048 → 2 classes |
| Loss function | Cross-entropy |
| Optimizer | AdamW |
| Learning rate | $5 \times 10^{-5}$ |
| Learning rate schedule | Cosine annealing (5-epoch warmup) |
| Batch size | 64 |
| Maximum epochs | 100 |
| Early stopping patience | 15 |
| Best model selection | Highest validation accuracy |
| Image size | 224×224 |
| Normalization | ImageNet mean and standard deviation |
| Training augmentations | Horizontal flip (p=0.5), rotation $\pm 10^\circ$, color jitter |

**Supplementary Table 3.** Performance of fine-tuned models across all three experiments: A) In-distribution, B) Out-of-distribution, and C) Initial weights. Values are reported as proportions with numerator/denominator fractions in parentheses. Bolded text demonstrates the highest performance.

| Model | Sensitivity | Specificity | PPV | NPV | Test Accuracy |
| --- | --- | --- | --- | --- | --- |
| **A.In-distribution** |  |  |  |  |  |
| Control | 0.66 (372/561) | **0.94 (946/1004)** | **0.87 (372/430)** | 0.83 (946/1135) | **0.84 (1318/1565)** |
| DINO | **0.68 (384/561)** | 0.92 (926/1004) | 0.83 (384/462) | **0.84 (926/1103)** | **0.84 (1310/1565)** |
| DINO-Ortho | 0.67 (376/561) | 0.93 (935/1004) | 0.84 (376/445) | 0.83 (935/1120) | **0.84 (1311/1565)** |
| **B.Out-of-distribution** |  |  |  |  |  |
| Control | 0.42 (710/1694) | **0.77 (2810/3671)** | **0.45 (710/1571)** | **0.74 (2810/3794)** | **0.66 (3520/5365)** |
| DINO | 0.35 (600/1694) | **0.77 (2824/3671)** | 0.41 (600/1447) | 0.72 (2824/3918) | 0.64 (3424/5365) |
| DINO-Ortho | **0.47 (793/1694)** | 0.66 (2419/3671) | 0.39 (793/2045) | 0.73 (2419/3320) | 0.60 (3212/5365) |
| **C.Initial weights** |  |  |  |  |  |
| Control | **0.86 (240/280)** | 0.92 (490/534) | 0.85 (240/284) | 0.92 (490/530) | 0.90 (730/814) |
| DINO | 0.85 (237/280) | **0.94 (504/534)** | **0.89 (237/267)** | **0.92 (504/547)** | **0.91 (741/814)** |
| DINO-Ortho | 0.85 (239/280) | 0.93 (496/534) | 0.86 (239/277) | 0.92 (496/537) | 0.90 (735/814) |

*PPV: positive predictive value; NPV: negative predictive value*

**Supplementary Table 4.** Qualitative ranking results, initial weights experiment (N = 50 calcaneus fractures). Individual rankings ranged from 1 (best) to 3 (worst). Bolded text represents the best average ranking.

| Model | Mean Rank |
| --- | --- |
| Control | 2.20* |
| DINO | 1.92* |
| DINO-Ortho | **1.89\*** |

\*Friedman test: $\chi^2(2) = 2.31$ , $P = .32$

**Supplementary Table 5.** Reliability metrics for qualitative ranking, initial weights experiment (N = 50 calcaneus fractures).

| Comparison | Weighted<br>kappa |
| --- | --- |
| Intra-rater, Rater 1 | 0.84 |
| Intra-rater, Rater 2 | 0.88 |
| Inter-rater (mean across sets) | 0.72 |

## References

1. Bhargavan M, Sunshine JH. Utilization of Radiology Services in the United States: Levels and Trends in Modalities, Regions, and Populations. Radiology. 2005;234(3):824–832. doi: 10.1148/radiol.2343031536.

2. York T, Franklin C, Reynolds K, et al. Reporting errors in plain radiographs for lower limb trauma—a systematic review and meta-analysis. Skeletal Radiol. 2022;51(1):171–182. doi: 10.1007/s00256-021-03821-9.

3. Robinson PJ, Wilson D, Coral A, Murphy A, Verow P. Variation between experienced observers in the interpretation of accident and emergency radiographs. Br J Radiol. 1999;72(856):323–330. doi: 10.1259/bjr.72.856.10474490.

4. Pinto A, Berritto D, Russo A, et al. Traumatic fractures in adults: missed diagnosis on plain radiographs in the Emergency Department. Acta Biomed Atenei Parm. 2018;89(1-S):111–123. doi: 10.23750/abm.v89i1-S.7015.

5. Lindsey R, Daluiski A, Chopra S, et al. Deep neural network improves fracture detection by clinicians. Proc Natl Acad Sci. 2018;115(45):11591–11596. doi: 10.1073/pnas.1806905115.

6. Guan B, Yao J, Zhang G, Wang X. Thigh fracture detection using deep learning method based on new dilated convolutional feature pyramid network. Pattern Recognit Lett. 2019;125:521–526. doi: 10.1016/j.patrec.2019.06.015.

7. Mongan J, Moy L, Kahn CE. Checklist for Artificial Intelligence in Medical Imaging (CLAIM): A Guide for Authors and Reviewers. Radiol Artif Intell. 2020;2(2):e200029. doi: 10.1148/ryai.2020200029.

8. Jones RM, Sharma A, Hotchkiss R, et al. Assessment of a deep-learning system for fracture detection in musculoskeletal radiographs. Npj Digit Med. 2020;3(1):144. doi: 10.1038/s41746-020-00352-w.

9. Zech JR, Badgeley MA, Liu M, Costa AB, Titano JJ, Oermann EK. Variable generalization performance of a deep learning model to detect pneumonia in chest radiographs: A cross-sectional study. Sheikh A, editor. PLOS Med. 2018;15(11):e1002683. doi: 10.1371/journal.pmed.1002683.

10. Banerjee I, Bhattacharjee K, Burns JL, et al. “Shortcuts” Causing Bias in Radiology Artificial Intelligence: Causes, Evaluation, and Mitigation. J Am Coll Radiol. 2023;20(9):842–851. doi: 10.1016/j.jacr.2023.06.025.

11. Litjens G, Kooi T, Bejnordi BE, et al. A survey on deep learning in medical image analysis. Med Image Anal. 2017;42:60–88. doi: 10.1016/j.media.2017.07.005.

12. Kandel I, Castelli M, Popovič A. Musculoskeletal Images Classification for Detection of Fractures Using Transfer Learning. J Imaging. 2020;6(11):127. doi: 10.3390/jimaging6110127.

13. Jing L, Tian Y. Self-Supervised Visual Feature Learning With Deep Neural Networks: A Survey. IEEE Trans Pattern Anal Mach Intell. 2021;43(11):4037–4058. doi: 10.1109/TPAMI.2020.2992393.

14. Huang S-C, Pareek A, Jensen M, Lungren MP, Yeung S, Chaudhari AS. Self-supervised learning for medical image classification: a systematic review and implementation guidelines. Npj Digit Med. 2023;6(1):74. doi: 10.1038/s41746-023-00811-0.

15. Caron M, Touvron H, Misra I, et al. Emerging Properties in Self-Supervised Vision Transformers. 2021. p. 9650–9660. https://openaccess.thecvf.com/content/ICCV2021/html/Caron_Emerging_Properties_in_Self-Supervised_Vision_Transformers_ICCV_2021_paper.html. Accessed January 14, 2026.

16. Tejani AS, Klontzas ME, Gatti AA, et al. Checklist for Artificial Intelligence in Medical Imaging (CLAIM): 2024 Update. Radiol Artif Intell. 2024;6(4):e240300. doi: 10.1148/ryai.240300.

17. Rajpurkar P, Irvin J, Bagul A, et al. MURA: Large Dataset for Abnormality Detection in Musculoskeletal Radiographs. arXiv; 2017. doi: 10.48550/ARXIV.1712.06957.

18. Abedeen I, Rahman MdA, Zohra Prottyasha F, Ahmed T, Mohmud Chowdhury T, Shatabda S. FracAtlas: A Dataset for Fracture Classification, Localization and Segmentation of Musculoskeletal Radiographs. figshare; 2023. p. 338412750 Bytes. doi: 10.6084/M9.FIGSHARE.22363012.

19. Selvaraju RR, Cogswell M, Das A, Vedantam R, Parikh D, Batra D. Grad-CAM: Visual Explanations from Deep Networks via Gradient-based Localization. arXiv; 2016; doi: 10.48550/ARXIV.1610.02391.

20. Rong Y, Leemann T, Borisov V, Kasneci G, Kasneci E. A Consistent and Efficient Evaluation Strategy for Attribution Methods. Int Conf Mach Learn. PMLR; 2022. p. 18770–18795. doi: 10.48550/ARXIV.2202.00449.

21. Landis JR, Koch GG. The measurement of observer agreement for categorical data. Biometrics. 1977;33(1):159–174.

22. Adebayo J, Gilmer J, Muelly M, Goodfellow I, Hardt M, Kim B. Sanity Checks for Saliency Maps. Adv Neural Inf Process Syst. 2018;31. https://papers.nips.cc/paper_files/paper/2018/hash/294a8ed24b1ad22ec2e7efea049b8737-Abstract.html. Accessed January 14, 2026.

23. Suara S, Jha A, Sinha P, Sekh AA. Is Grad-CAM Explainable in Medical Images? arXiv; 2023; doi: 10.48550/ARXIV.2307.10506.

24. Azizi S, Mustafa B, Ryan F, et al. Big Self-Supervised Models Advance Medical Image Classification. 2021. p. 3478–3488. https://openaccess.thecvf.com/content/ICCV2021/html/Azizi_Big_Self-Supervised_Models_Advance_Medical_Image_Classification_ICCV_2021_paper.html. Accessed January 14, 2026.

25. Wang X, Yang S, Zhang J, et al. Transformer-based unsupervised contrastive learning for histopathological image classification. Med Image Anal. 2022;81:102559. doi: 10.1016/j.media.2022.102559.

26. Holmberg OG, Köhler ND, Martins T, et al. Self-supervised retinal thickness prediction enables deep learning from unlabelled data to boost classification of diabetic retinopathy. Nat Mach Intell. 2020;2(11):719–726. doi: 10.1038/s42256-020-00247-1.

